# Investigation on the clinicopathological characteristics and BRCA1/2 gene variation of patients in breast cancer pedigrees in Northern Henan Province

**DOI:** 10.1101/2023.05.25.23290419

**Authors:** Hui Zhao, Lei Wang, Yueqing Feng, Junzheng Yang

## Abstract

**Objectives:** To investigate the clinicopathological characteristics of breast cancer patients and the pathogenic gene variation of BRCA1/2 in breast cancer pedigrees in Northern Henan province, to provide the evidences for treatment or prevention of breast cancer.

**Methods:** 214 breast cancer patients from different families admitted to Xinxiang Central Hospital/Fourth Clinical College of Xinxiang Medical University from November 2018 to January 2021 were selected, DNA samples were extracted from patient and the exon and intron splicing regions in the coding sequences of BRCA1 and BRCA2 genes were amplified by PCR, the amplified products were screened and the abnormal segments were confirmed by Sanger sequencing; finally, Integrative Genomics Viewer software and Codon Code Aligner software was used to verify the candidate pathogenic sites in breast cancer pedigrees.

**Results:** Among 214 cases of breast cancer patients, there were 177 patients with unilateral breast cancer and 37 patients with bilateral breast cancer, accounting for 82.71% and 17.29% in 214 breast cancer patients, respectively; there were 122 patients in premenopausal menstruation and 92 patients in postmenopausal state at the time of diagnosis, accounting for 57.01% and 42.99% in 214 breast cancer patients, respectively; there were 137 patients with the tumor diameter at 2-5 cm, 60 patients with tumor diameter≤2 cm, 17 patients with tumor diameter>5 cm, accounting for 64.02%, 28.04%, and 7.94%, respectively; there were 183 patients were invasive ductal carcinoma, accounting for 85.51% in 214 breast cancer patients; regional lymph node metastasis was mainly negative (130 cases, accounting for 60.75% in 214 breast cancer patients), TNM staging was mainly stage II (138 cases, accounting for 64.49% in 214 breast cancer patients), and histological classification was mainly stage II. The gene sequencing results demonstrated that a total of 20 pathogenic mutations were found including 17 BRCA1 gene mutations and 3 BRCA2 gene mutations in 214 patients with familial breast cancer; there were 11 frameshift mutations, 3 nonsense mutations and 3 splice mutations in 17 cases BRCA1 gene mutation, and all 3 BRCA2 gene mutations were frameshift mutations; especially, the 1100delT site mutation of BRCA1 gene was repeated in 3 patients with breast cancer. 18 high frequency SNP (frequency ≥ 5%) loci were found in 214 breast cancer patients, there were 17 the mutation frequency was higher than that of the normal population; especially, the mutation frequency of rs799917 is lower than that of normal population. Finally, we analyzed the clinical characteristics of rs80356892 polymorphism carriers in 214 breast cancer patients, found that there were 5 patients with rs80356892 mutation, including 3 patients with bilateral primary breast cancer, accounting for 60%, and the 5 patients with rs80356892 mutation had the family history of disease.

**Conclusion:** the clinicopathological characteristics and BRCA1/2 gene variation of patients in breast cancer pedigrees in Northern Henan Province had certain specificity and regional characteristics, these data may provide some useful information for prevention or treatment for breast cancer.

## Introduction

It has been reported that breast cancer is becoming one of the malignant diseases with high incidence in the world, accounting for about 12% in all kinds of female cancers^[1]^, and data demonstrated that 5-10% of breast cancer is hereditary because of genetic susceptibility^[2,3]^. up to now, the genes found susceptible to breast cancer account for 25-30% in hereditary breast cancers^[4]^; interestingly, a large number of patients with breast cancer family history do not carry risk variations in these genes, indicating that there may be other genetic risk factors^[5]^. In clinic, identifying new susceptible genes and missing heritability in breast cancer will help us to improve the genetic risk estimation for the accuracy of population, and to find ways to translate these findings into clinical practice, to carry out early detection and prevention in identifying individuals at risk. It has been reported that BRCA1/2 gene is one kind of gene regulation factors and the most important tumor-related genes which will result in the high risk and high mortality of the breast cancer, and it also is the two main susceptible genes of hereditary breast cancer^[6,7,8]^. Therefore, to investigate the pathogenic BRCA1/2 gene variation in breast cancer pedigrees has important clinical significance. For this purpose, we investigated the clinicopathological characteristics of breast cancer patients, and analyzed the BRCA1 and BRCA2 gene mutations and clinical characteristics of regional familial breast cancer patients and their healthy female relatives, which may provide evidences and new ideas for future gene diagnosis and gene therapy of breast cancer.

## 1 Materials and methods

### 1.1 General information

214 breast cancer patients from different families were collected from Xinxiang Central Hospital/Fourth Clinical College of Xinxiang Medical University from November 2018 to January 2021, there were 133 families with familial breast cancer were screened; at least one breast cancer patient and one family member were selected from each breast cancer family. A total of 214 samples including 162 breast cancer patients and 52 family members were collected, and all the subjects were Chinese.

### 1.2 Inclusion criteria

(1) At least two generations of immediate family members have breast cancer patients; (2) the age of onset is≤40 years old; (3) two or more patients was identified as bilateral breast cancer in the same generation; (4) patients with complete examination data; (5) being in non-pregnancy and lactation period.

### 1.3 Exclusion criteria

(1) Patients with history of malignant tumor; (2) standardized comprehensive therapy for breast cancer has been adopted; (3) patients with severe liver and kidney dysfunction; (4) patients who were unwilling to participate in the study; (5) people who with mental disorders.

### 1.4 Methods

Clinicopathological information of familial breast cancer patients including patient’s disease type, menstrual status, tumor size, regional lymph node metastasis, pathological type, TNM stage, histological classification at the time of diagnosis.

Genomic DNA and peripheral blood DNA were extracted from tumor tissue and peripheral blood samples of patients (DNA rapid extraction kit). Subsequently, the genomic DNA library was constructed (HTP/LTP preparation kit, KAPA). For Sanger sequencing analysis, DNA was extracted from the patient’s peripheral blood samples, and then polymerase chain reaction PCR amplification (Q5 High-Fidelity DNA Polymerase, NEB) (forward primer sequence: TGGGAGAGTCATGCATCTCA; reverse primer sequence: TCGGTACCCTGAGCCAAAT) and PCR products were purified, the BRCA1 and BRCA2 gene were analyzed using the Integrated Genomics Viewer software and the Codon Code Aligner, respectively. The 49 exon sequences of BRCA1 and BRCA2 coding regions in patients’ peripheral blood were detected and analyzed, SNP was used to detect the mutation frequency of BRCA1/2 gene in breast cancer patients.

## 2 Results

### 2.1 The clinicopathological information of familial breast cancer patients

Among 214 cases of breast cancer patients, there were 177 patients were unilateral cancer, accounting for 82.71% in the total of 214 breast cancer patients; 37 patients with bilateral cancer, accounting for 17.29% in the total of 214 breast cancer patients; the number of patients with premenopausal menstruation at the time of diagnosis was 122, accounting for 57.01% in the total of 214 breast cancer patients; there were 92 patients in postmenopausal state, accounting for 42.99% in the total of 214 breast cancer patients; 137 cases patients with the tumor size were 2-5 cm, accounting for 64.02% in the total of 214 breast cancer patients; 60 patients with tumor diameter ≤ 2 cm, accounting for 28.04% in the total of 214 breast cancer patients; 17 patients with tumor diameter>5 cm, accounting for 7.94% in the total of 214 breast cancer patients; compared from the perspective of pathological type, the most common histological type of tumor is invasive ductal carcinoma (183 cases), accounting for 85.51% in the total of 214 breast cancer patients, regional lymph node metastasis was mainly negative (130 cases, accounting for 60.75% in the total of 214 breast cancer patients), TNM staging was mainly stage II (138 cases, accounting for 64.49% in the total of 214 breast cancer patients), and histological classification was mainly stage II (Table 1).

**Table 1.** The clinicopathological information of familial breast cancer patients.

|  | Breast cancer type |  | Menstrual status |  | Tumor size/cm |  |  |
| --- | --- | --- | --- | --- | --- | --- | --- |
| | unilateral | bilateral | pre | post | $\leq 2$ | 2-5 | $>5$ |
| Number/n | 177 | 37 | 122 | 92 | 60 | 137 | 17 |
| Percentage/% | 82.71 | 17.29 | 57.01 | 42.99 | 28.04 | 64.02 | 7.94 |

|  | Regional lymph node metastasis |  | Pathological type |  | TNM stage |  |  | Histological classification |  |  |
| --- | --- | --- | --- | --- | --- | --- | --- | --- | --- | --- |
|  | Negative | Positive | Invasive ductal carcinoma | Non-invasive ductal carcinoma | 0+I | II | III | I | II | III |
| Number/n | 130 | 84 | 183 | 31 | 61 | 138 | 15 | 31 | 145 | 38 |
| Percentage/% | 60.75 | 39.25 | 85.51 | 14.49 | 28.5 | 64.49 | 7.01 | 14.48 | 67.76 | 17.76 |

### 2.2 Gene sequencing analyzed the BRCA1 and BRCA2 gene mutations in 214 breast cancer patients

Among 214 familial breast cancer patients, 20 patients had BRCA1 or BRCA2 gene mutations, the mutation rate was 9.35%. 17 patients had BRCA1 gene mutations, accounting for 85% in the total 20 gene mutations; 3 patients had BRCA2 gene mutations, accounting for 15% in the total 20 gene mutations; among 17 BRCA1 gene mutations, there were 11 frameshift mutations, 3 splice mutations and 3 nonsense mutations; all the BRCA2 gene mutations were frameshift mutations (Table 2).

**Table 2.**
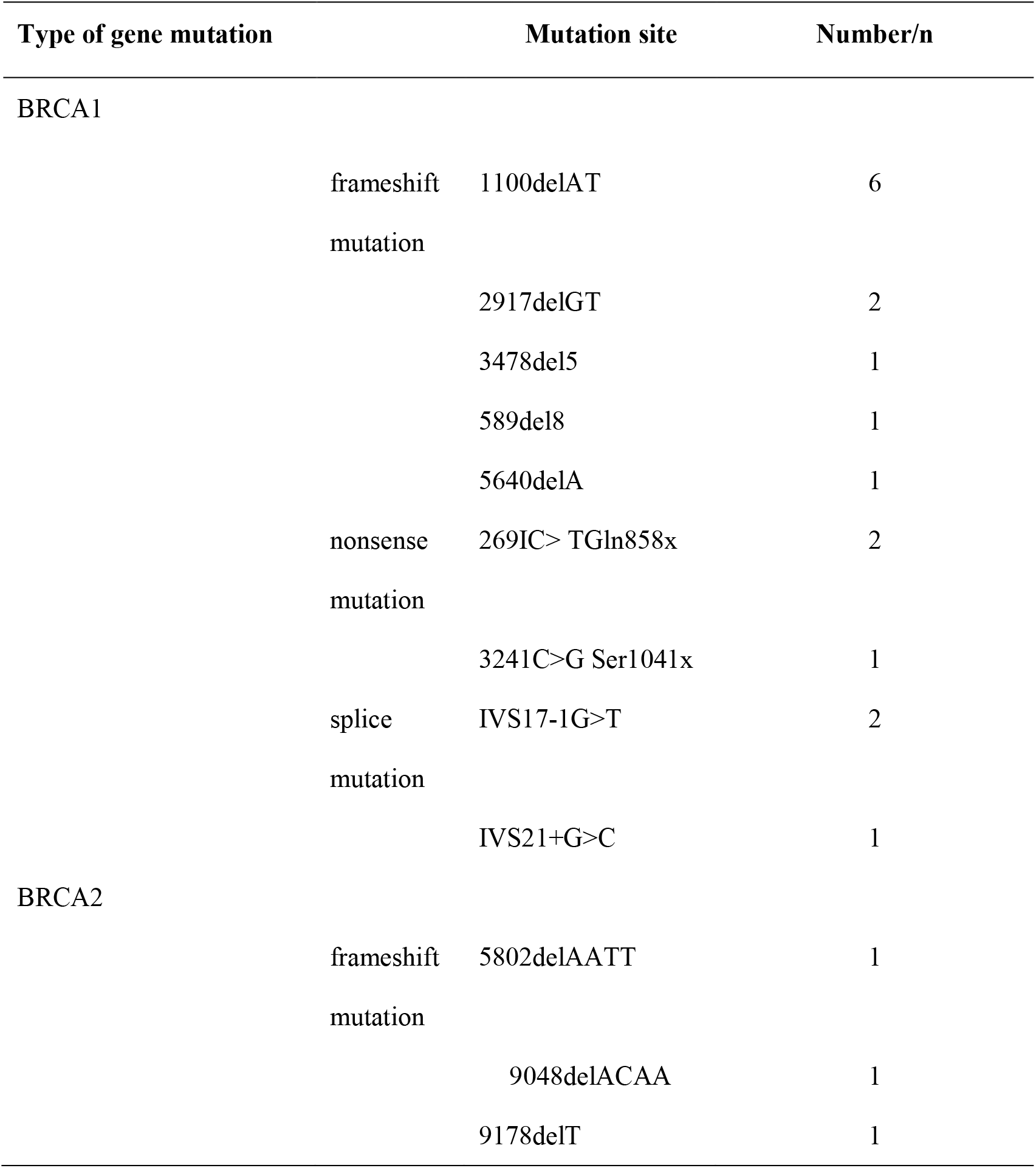
Analysis of BRCA1 and BRCA2 gene mutations in 214 familial breast cancer patients.

### 2.3 Compared the SNP mutation frequency of BRCA1/2 gene in BC patients and the normal population

In 214 patients with breast cancer, there were 34 mutations were found at the SNP site of BRCA1/2 gene (the mutation rate was 15.89%), there were 14 mutations were located in BRCA1 and 20 mutations were located in BRCA2; 18 high frequency SNP (frequency≥5%) loci were found, and there was 17 high frequency SNP loci in BC patients was higher than that in the normal population; especially, the mutation frequency site rs799917 in BC patients was lower than that in the normal population (mutation frequency in BC patients/mutation frequency in normal polulation=51.3/54.6) (Table 3).

**Table 3.**
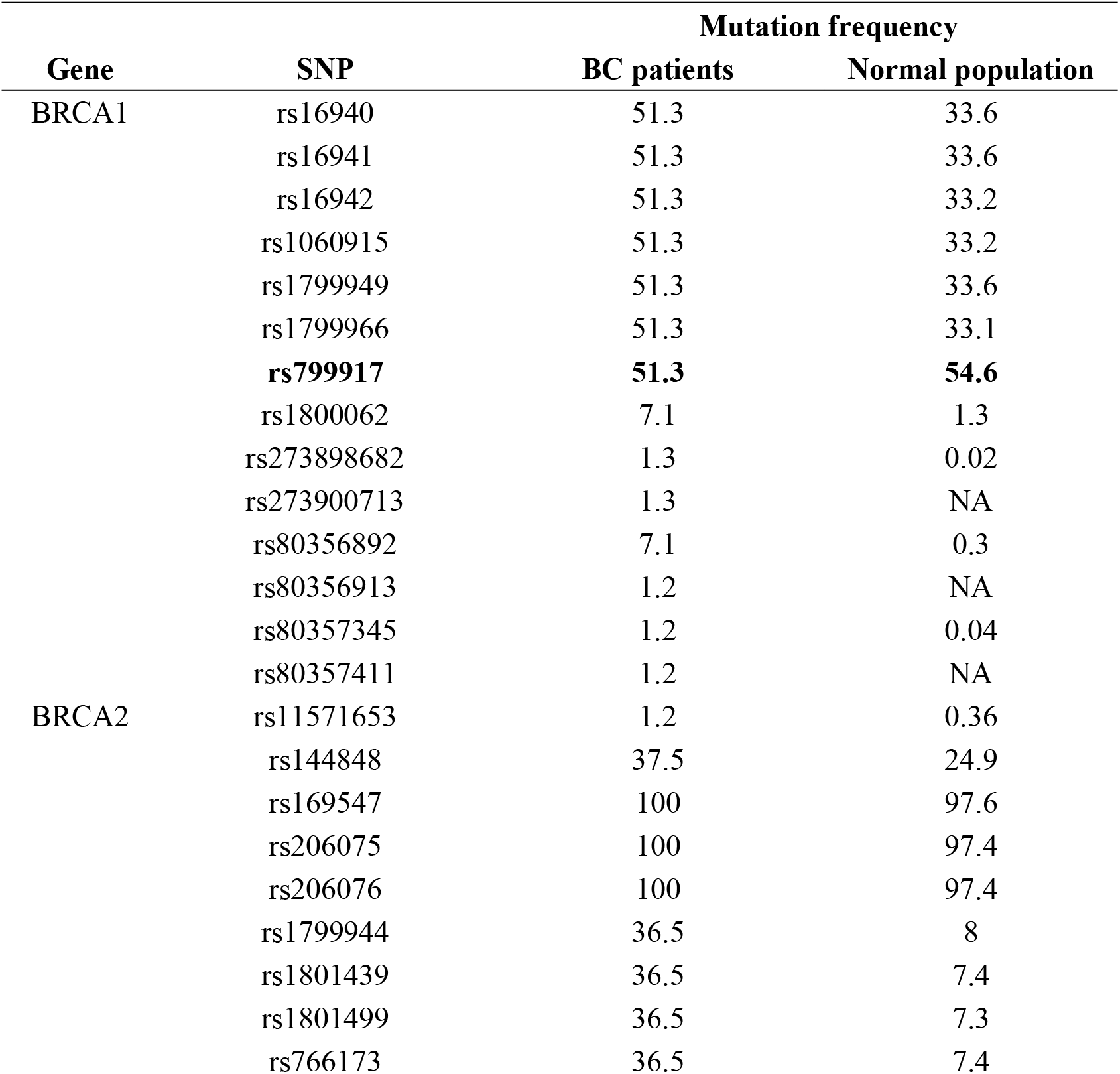
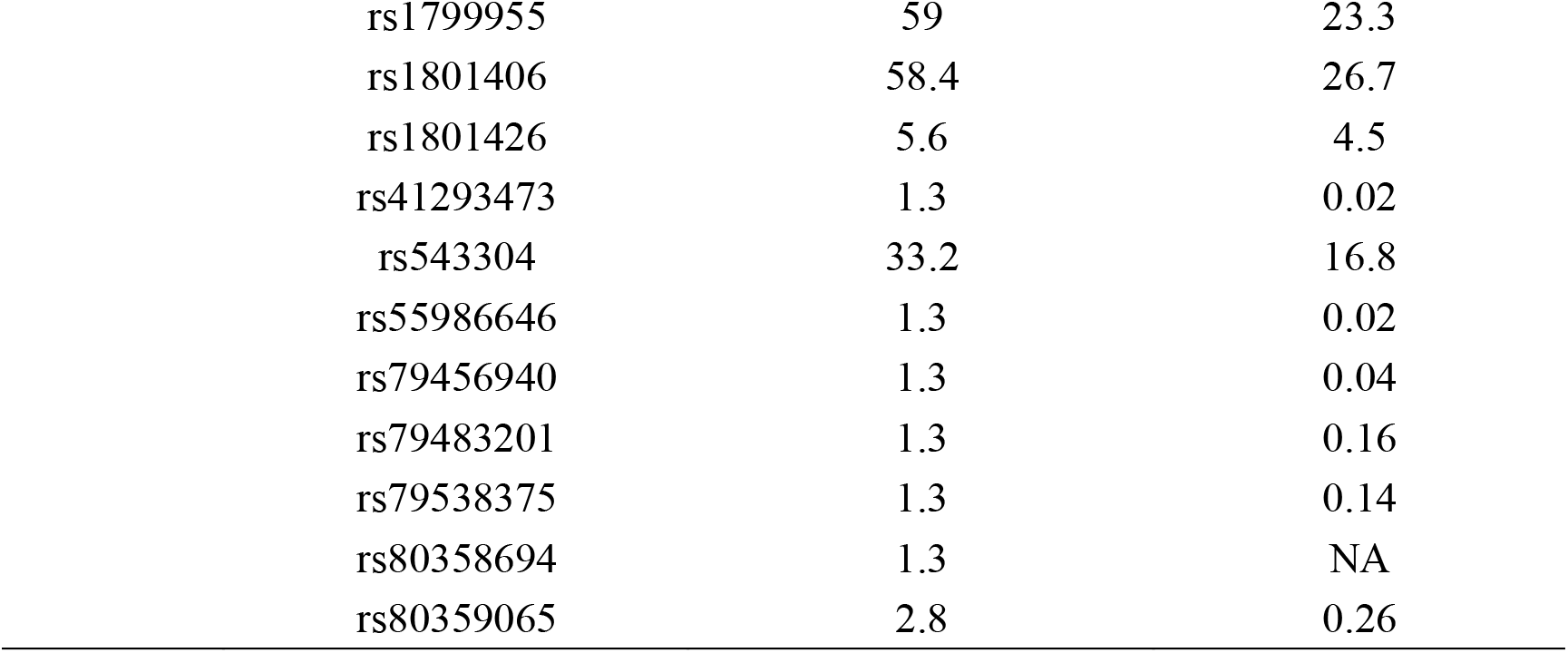
Comparison of mutation frequency site in BRCA1/2 gene in BC patients and normal population.

| Gene | SNP | Mutation frequency |  |
| --- | --- | --- | --- |
|  |  | BC patients | Normal population |
| BRCA1 | rs16940 | 51.3 | 33.6 |
|  | rs16941 | 51.3 | 33.6 |
|  | rs16942 | 51.3 | 33.2 |
|  | rs1060915 | 51.3 | 33.2 |
|  | rs1799949 | 51.3 | 33.6 |
|  | rs1799966 | 51.3 | 33.1 |
|  | **rs799917** | **51.3** | **54.6** |
|  | rs1800062 | 7.1 | 1.3 |
|  | rs273898682 | 1.3 | 0.02 |
|  | rs273900713 | 1.3 | NA |
|  | rs80356892 | 7.1 | 0.3 |
|  | rs80356913 | 1.2 | NA |
|  | rs80357345 | 1.2 | 0.04 |
|  | rs80357411 | 1.2 | NA |
| BRCA2 | rs11571653 | 1.2 | 0.36 |
|  | rs144848 | 37.5 | 24.9 |
|  | rs169547 | 100 | 97.6 |
|  | rs206075 | 100 | 97.4 |
|  | rs206076 | 100 | 97.4 |
|  | rs1799944 | 36.5 | 8 |
|  | rs1801439 | 36.5 | 7.4 |
|  | rs1801499 | 36.5 | 7.3 |
|  | rs766173 | 36.5 | 7.4 |

|  |  |  |
| --- | --- | --- |
| rs1799955 | 59 | 23.3 |
| rs1801406 | 58.4 | 26.7 |
| rs1801426 | 5.6 | 4.5 |
| rs41293473 | 1.3 | 0.02 |
| rs543304 | 33.2 | 16.8 |
| rs55986646 | 1.3 | 0.02 |
| rs79456940 | 1.3 | 0.04 |
| rs79483201 | 1.3 | 0.16 |
| rs79538375 | 1.3 | 0.14 |
| rs80358694 | 1.3 | NA |
| rs80359065 | 2.8 | 0.26 |

### 2.4 The clinical characteristics of rs80356892 polymorphism carriers

The result of the clinical characteristics of rs80356892 polymorphism carriers demonstrated that there were 5 patients with rs80356892 mutation, including 3 patients with bilateral primary breast cancer, accounting for 60%, and all the 5 patients with rs80356892 mutation had the family history of disease (Table 4).

**Table 4.** The clinical characteristics of rs80356892 site polymorphism carriers.

| No. | Type of breast cancer and age | Family history |
| --- | --- | --- |
| 1 | BC, 36-40 years old | Sister, BBC, 33-38 years old |
| 2 | BC, 30-35 years old | Mother, BC, 44-50 years old |
| 3 | BBC, 35-40 years old | Maternal aunt, X,<br>45-55 years old |
| 4 | BBC, 33-38 years old | Sister, BC, 35-40 years old |
| 5 | BBC, 41-49 years old | Maternal cousin, BC, 35-40<br>years old |

Abbreviation: BC, breast cancer; BBC, bilateral breast cancer.

## Discussion

Breast cancer is a kind of diseases that seriously affects women’s health and quality of life no matter in developed countries or developing countries. The clinicopathological characteristics in different countries and regions play the important role in prevention and treatment of breast cancer. In our data, we found that unilateral cancer was the main type of breast cancer in the northern Henan Province; in the total of 214 breast cancer patients, there were 64.02% patients with the tumor diameter at 2-5cm, 28.04% patients were tumor diameter≤2cm, 7.94% patients with tumor diameter>5cm; the most common histological type of breast cancer was invasive ductal carcinoma; regional lymph node metastasis was mainly negative (130 cases, accounting for 60.75% in the total of 214 breast cancer patients); TNM staging was mainly stage II (138 cases, accounting for 64.49% in the total of 214 breast cancer patients), and histological classification was mainly stage II in the total of 214 breast cancer patients. Those data demonstrated that the clinicopathological characteristics of breast cancer in Northern Henan province had the certain characteristics, which may provide the guidance for clinical staff.

BRCA1 and BRCA2 play an important role in repairing DNA double strand breaks through homologous recombination. BRCA mutations lead to BRCA protein dysfunction, furtherly result in DNA repair errors and genetic aberrations ^[9,10]^. The evidences have demonstrated that BRCA1 and BRCA2 mutations are positively correlated with the increased risk of hereditary breast and ovarian cancer syndrome (HOBC), if there was the gene mutation of BRCA1 or BRCA2, the incidence rates of breast cancer and ovarian cancer before the age of 70 years old will be higher, and it has been reported that the number of individuals with cancer in the family was generally considered as a strong clinical predictor of HBOC ^[11,12,13,14,15]^. In our article, we also found the frameshift mutation in BRCA1 gene; the results of SNP mutation frequency of BRCA1/2 gene and the clinical characteristics of rs80356892 polymorphism carriers demonstrated that there were 15.89% breast cancer patients had found the SNP site mutations at BRCA1/2 gene in the total of 214 breast cancer patients; there were 14 mutations were located in BRCA1 and 20 mutations were located in BRCA2; especially, there were 5 patients with rs80356892 mutation and with the family history of disease. Those data may provide some reference information for clinic staff.

## Conclusion

The clinicopathological characteristics, BRCA1 and BRCA2 gene mutation frequency and BRCA1 and BRCA2 gene mutation loci in breast cancer patients in Northern Henan Province has the unique distribution characteristics, those data may provide the clinical guidance significance for prevention and treatment of breast cancer.

## Data Availability

The authors confirm that the data supporting the findings of this study are available within the article [and/or] its supplementary materials.

## Conflicts of interests

The authors declared that there are no conflicts of interests in this article.

## Funding

None.

## Acknowledgments

None.

## Notes

### Competing Interest Statement

The authors have declared no competing interest.

### Funding Statement

This study did not receive any funding

